# Centenarians maintain cognition by resisting amyloid-β or decoupling it from tau propagation

**DOI:** 10.64898/2026.09.18.26363376

**Authors:** Susan K. Rohde, Maruelle C. Luimes, Annemieke J.M. Rozemuller, Marc Hulsman, Sven J. van der Lee, Sietske A.M. Sikkes, Timothy E. Richardson, Jeroen J.M. Hoozemans, Jamie M. Walker, Henne Holstege

## Abstract

Centenarians who maintain cognitive health provide a unique opportunity to investigate naturally occurring mechanisms that protect against Alzheimer’s disease. We examined the relationship between subregional amyloid-beta and p-tau distributions in the medial temporal lobe and cognitive performance in 112 centenarians. We show that amyloid-beta pathology was associated with increased parahippocampal, but not hippocampal, p-tau burden and lower cognitive performance. However, the majority of the centenarians maintained high cognitive performance until death, and they appeared to be protected against cognitive decline through two distinct mechanisms: 44% were resistant to amyloid-beta accumulation, while 28% harbored substantial amyloid-beta pathology but resisted downstream p-tau propagation. These findings suggest the existence of natural resilience mechanisms that uncouple amyloid-beta pathology from driving downstream pathogenic tau progression. Both mechanisms warrant further exploration, as they may offer complementary therapeutic entry-points: preventing the accumulation of Aβ pathology altogether, or limiting downstream p-tau propagation once Aβ pathology has already emerged.

## Introduction

Alzheimer’s disease (AD) is the most common cause of dementia, affecting an estimated 50 million people worldwide^1^. Neuropathologically, AD is defined by amyloid-beta (Aβ) plaques and neurofibrillary tangles (NFT) composed of hyperphosphorylated tau (p-tau). The prevalence of both AD-related cognitive decline and AD neuropathologic change (ADNC) become increasingly common with advancing age^2–4^. However, some individuals reach extreme old age (>100 years) without cognitive decline and have variable levels of Aβ and p-tau pathology^5,6^. These cognitively healthy centenarians constitute a unique cohort for investigating the natural protective mechanisms that underly maintaining a healthy functioning brain until extreme old age. We recently demonstrated that in a unique cohort of 95 self-reported cognitively healthy centenarians from the Dutch 100-plus Study, two-thirds accumulated no or little neocortical Aβ burden, which was associated with maintaining high levels of cognitive performance until death. On the other hand, one-third had neocortical Aβ burden comparable to that observed in AD patients, which associated with lower executive functioning^7^. This aligns with the amyloid-cascade hypothesis, which proposes Aβ accumulation as a trigger for downstream p-tau pathology, neurodegeneration, and cognitive decline^8^. Strikingly, several centenarians maintained high cognitive performance despite having high Aβ burden, suggesting that they were resilient to the accumulated Aβ pathology. These centenarians showed unexpectedly low p-tau spreading, with lower Braak NFT stages compared to individuals with similarly high Aβ burden and impaired cognition^7^.

Previous studies have demonstrated that Aβ influences the spatial distribution of p-tau within the medial temporal lobe (MTL)^9–12^, which includes the hippocampus. In AD, where Aβ accumulation is abundant, p-tau predominantly affects the subiculum and cornu ammonis 1 (CA1) subregions of the hippocampus. In contrast, in primary age-related tauopathy (PART), which is characterized by p-tau pathology in the MTL in absence of substantial Aβ pathology^13^, p-tau is initially more concentrated in CA2, resulting in a higher CA2/CA1 p-tau burden ratio^10^. PART is observed in ∼20-30% of individuals aged >80^14^ and in ∼40% of those aged >90^15^, and is generally associated with less cognitive impairment than p-tau in the presence of Aβ (i.e., AD)^13^. Altogether, this leads us to question whether some centenarians have an altered relationship between Aβ and p-tau distribution and burden, which could point to endogenous mechanisms of resilience.

Here, we assessed 112 post-mortem centenarian brains for Aβ and p-tau pathology and grouped them as Aβ-low (i.e., PART) or Aβ-high (i.e., ADNC), according to the golden-standard neuropathological criteria^13,16^. Moreover, we assessed Aβ and p-tau pathological burden across nine MTL subregions and examined its association with ante-mortem cognitive performance. Interestingly, 44% of centenarians were resistant to Aβ pathology and met the neuropathological criteria for PART^13^. Of the centenarians with high Aβ pathology (56%), half had p-tau distributions in the MTL comparable to AD patients, and these centenarians were more likely to have lower cognition (28% of the entire cohort). This is in support of the amyloid-cascade hypothesis. Interestingly, the other half of Aβ-high centenarians (28% of the entire cohort) resisted the accumulation of p-tau typical for AD, despite the presence of Aβ, and they maintained cognitive performance. This latter group of centenarians points to a natural mechanism of resilience in the cascade of Aβ driven tau pathology and cognitive decline.

## Results

### ADNC and PART in centenarians

The majority of the 112 centenarians were female (71%) and the median age at brain donation was 103.3 (IQR 102.3-104.8, min 100.4, max 111.8; **Table 1**). All centenarians had at least some p-tau pathology, with Braak NFT stages ranging between I-V (median III, IQR III-IV; **Table 1**). Of all centenarians, 56% had advanced spreading of Aβ (i.e., Thal phase ≥ 3, n=63) hereafter referred to as Aβ-high centenarians (**Figure 1A-B**), while 44% had no or limited spreading of Aβ (i.e., Thal phase ≤2, n=49), hereafter referred to as Aβ-low centenarians. Of the entire centenarian cohort, 9% were Aβ-low with definite PART (n=10, Thal 0, Braak I-IV, CERAD NP score 0), 35% were Aβ-low with possible PART (n=39, Thal 1-2, Braak I-IV, CERAD NP score 0-1), 10% were Aβ-high with low ADNC (n=11, Thal 3, Braak I-II, CERAD NP score 0-2), 40% were Aβ-high with intermediate ADNC (n=45, Thal 3-5, Braak III-V, CERAD NP score 0-3), and 6% were Aβ-high with high ADNC (n=7, Thal 4-5, Braak V, CERAD 2-3). Between these diagnostic groups, there were no significant differences in age (p=0.117), sex (p=0.466), hippocampal sclerosis (p=0.182), or Braak LB stage (p=0.48), but TDP-43 stage tended to be higher in Aβ-high centenarians, especially in those with intermediate and high ADNC, with borderline significance (p=0.06; **Figure S1**).

**Table 1.** Demographic and neuropathological characteristics of the centenarian (CEN) cohort, and the Alzheimer’s disease (AD) and primary age-related tauopathy (PART) reference cohorts.

| Cohort | N | Sex<br>(F:M %) | Median age at death<br>(Q1-Q3) [min-max] | Thal Phase<br>(0\|1\|2\|3\|4\|5) | Braak Stage<br>(I\|II\|III\|IV\|V\|VI) | CERAD NP<br>Score<br>(0\|1\|2\|3) |
| --- | --- | --- | --- | --- | --- | --- |
| CEN | 112 | 71:29 | 103.3 (102.3-104.8)<br>[100.4-111.8] | 10\|24\|15\|36\|16\|11 | 6\|16\|46\|36\|8\|0 | 52\|32\|24\|4 |
| AD | 11 | 36:64 | 84 (72-86) [53-100] | 0\|0\|0\|3\|1\|5 | 0\|0\|0\|0\|5\|3\|3 | 0\|2\|3\|6 |
| PART | 7 | 86:14 | 88 (78-92) [71-103] | 4\|2\|1\|0\|0\|0 | 0\|1\|3\|3\|0\|0\|0 | 7\|0\|0\|0 |

**Figure 1.**
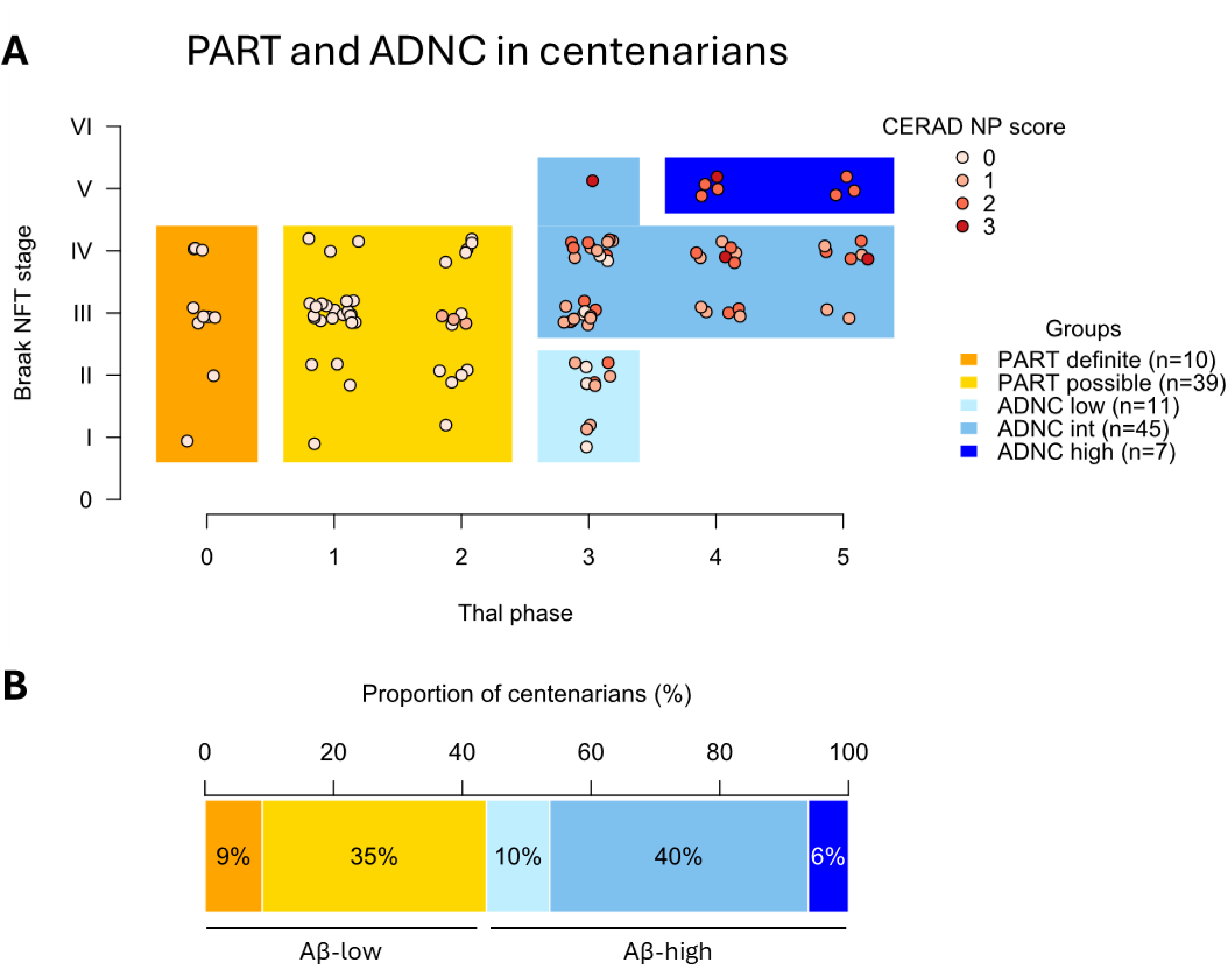
Alzheimer’s disease neuropathologic change (ADNC) and primary age-related tauopathy (PART) in centenarians. **(A)** The relation between Thal phase for amyloid-beta (Aβ), Braak stage for neurofibrillary tangles (NFT) and CERAD score for neuritic plaques (NP) in 112 centenarians evaluated in this study. Based on these three scores, centenarians could be classified as having primary age-related tauopathy (PART; definite or possible)^13^ or ADNC (low, intermediate (int) and high)^25^. **(B)** The percentage of centenarians meeting the criteria for the different subgroups of PART or ADNC.

### CA2/CA1 p-tau burden ratio does not differentiate between Aβ-low and Aβ-high centenarians

In a reference cohort of 7 Aβ-low PART cases (median age 88, IQR 78-92) and 11 Aβ-high AD cases (median age 84, IQR 72-86; **Table 1**), the median CA2/CA1 p-tau burden ratio was respectively 3.04 (range 1.6-4.2) versus 1.15 (range 0.8-1.4; p=0.005; **Figure 2A**). This clear distinction was not observed in the centenarian cohort, in which the median CA2/CA1 burden p-tau ratio was similar between Aβ-low and Aβ-high centenarians: 1.33 (n=49, range 0.3-3.6) versus 1.26 (n=63; range 0.2-4.3; p=0.263; **Figure 2A**). As hippocampal sclerosis (HS) could potentially decrease the apparent p-tau burden in CA1 and inflate the CA2/CA1 p-tau burden ratio, we ran a separate analysis excluding 23 centenarians with HS and still did not observe a difference in CA2/CA1 p-tau burden ratio between Aβ-low and Aβ-high centenarians (1.29 (n=41) vs 1.16 (n=48), p=0.242; **Figure S2**). In fact, a high CA2/CA1 p-tau burden ratio (>1.6, corresponding to the minimum ratio observed in the PART reference-cohort) was found in only 43% of Aβ-low centenarians (21/49) and as many as 33% of the Aβ-high centenarians (21/63; representative cases shown in **Figure 2B**). In addition, the CA2/CA1 p-tau burden ratio did not correlate with Aβ burden in any subregion or Thal phase within the centenarian cohort (**Figure 3**; **Figure S3**). Together, these findings indicate that in this centenarian cohort: 1) a PART-like p-tau distribution is not exclusively observed in Aβ-low individuals; and 2) not all centenarians with no/low Aβ pathology have a PART-like p-tau distribution.

**Figure 2.**
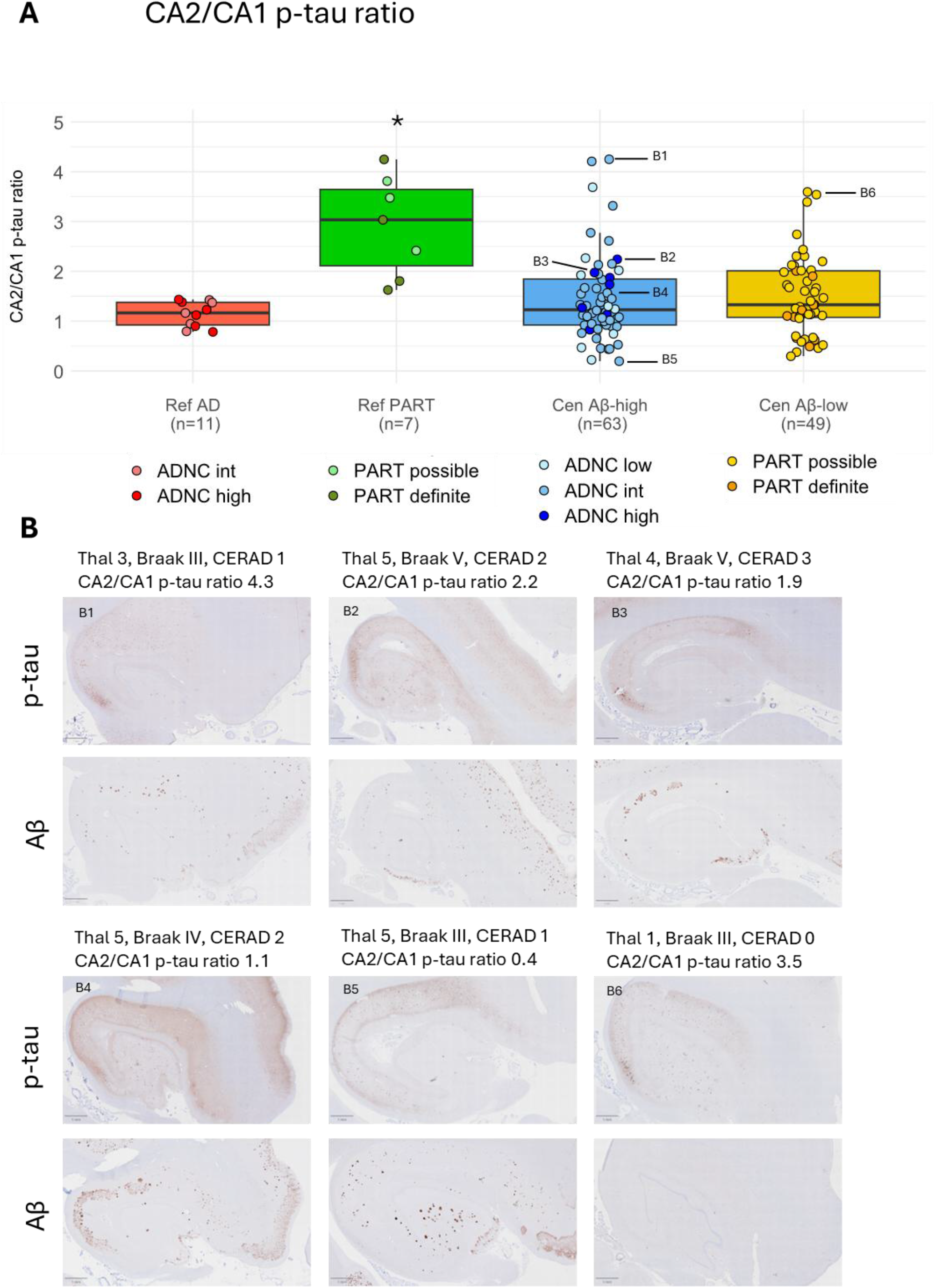
CA2/CA1 p-tau burden ratio comparable in Aβ-high and Aβ-low centenarians. **(A)** CA2/CA1 p-tau burden ratios compared between 11 clinicopathologically confirmed AD patients with Alzheimer’s disease neuropathologic change (ADNC) intermediate (int) or high, 7 primary age-related tauopathy (PART) cases for reference, and 63 Aβ-high centenarians (ADNC low, int, or high), and 49 Aβ-low centenarians (possible and definite PART; Figure 1). CA2/CA1 p-tau burden ratio was higher in the PART reference-cohort compared to all other groups (Kruskal-Wallis rank sum test with Dunn’s post-hoc test corrected for FDR; PAD=0.005; plADNC=0.0163; pADNC=0.0043; pPART=0.0071, indicated with *), but no other significant differences were identified. **(B)** Examples of hippocampal p-tau (AT8) and Aβ (6F/3D) pathology in three Aβ-high centenarians with a high CA2/CA1 p-tau burden ratio (B1-3), two Aβ-high centenarians with a low CA2/CA1 p-tau burden ratio (B4-5), and one Aβ-low centenarians with a high CA2/CA1 p-tau burden ratio (B6).

**Figure 3.**
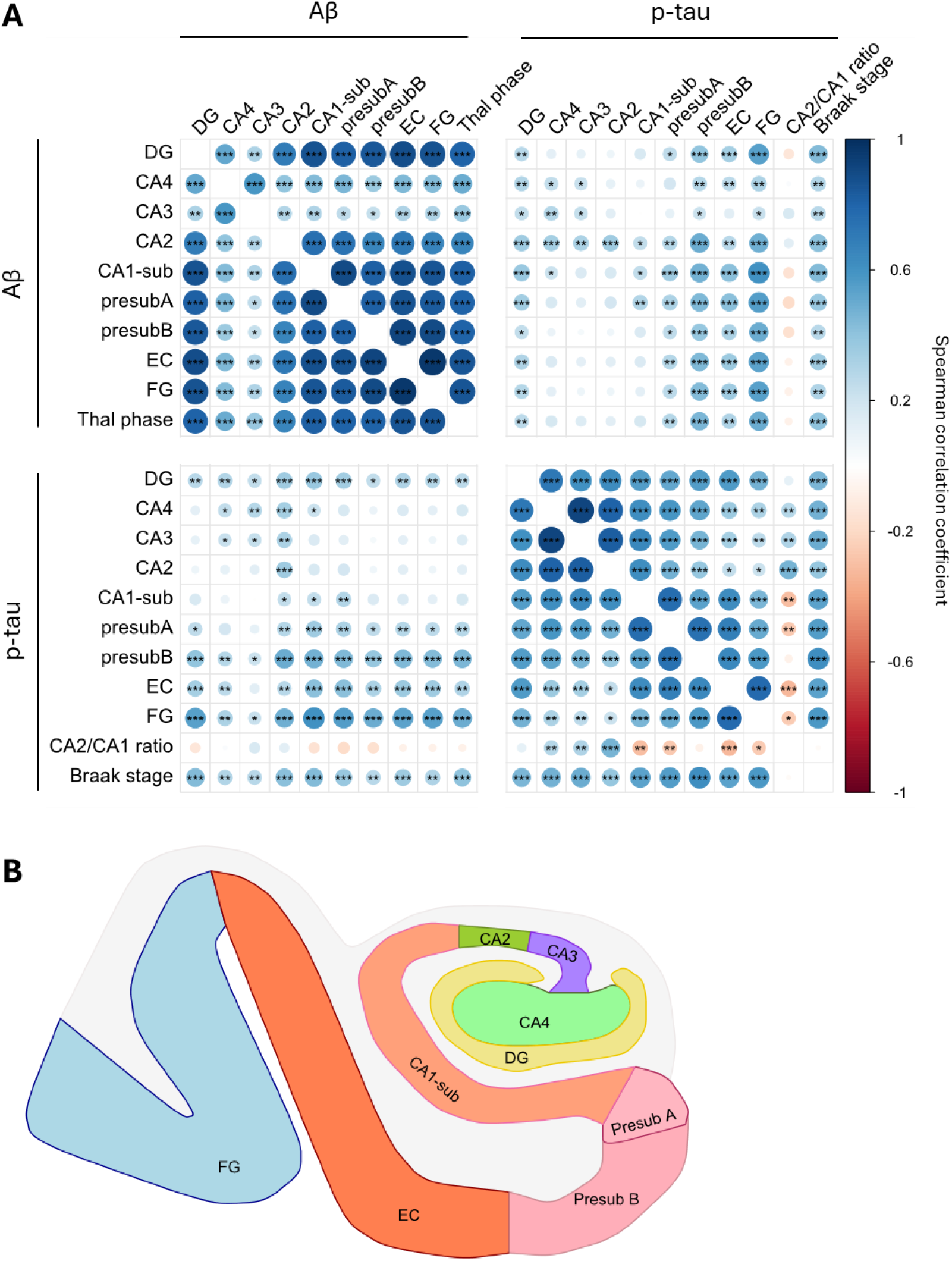
Correlation between subregional amyloid-beta burden (Aβ) and hyperphosphorylated tau (p-tau) burden. **(A)** Correlations between Thal phase, Braak stage, and Aβ and p-tau burden in nine subregions (illustrated in B) in the medial temporal lobe in 112 centenarians. Color and size of the dots indicate Spearman correlation coefficient and p (* ≤0.05 and ** ≤0.01, ***≤0.001) corrected for false discovery rates (FDR) using Benjamini & Hochberg method. **(B)** Illustration of the subregions analyzed: dentate gyrus (DG); cornu ammonis (CA)4-2; CA1-subiculum (CA1-sub); presubiculum A and B (presubA; presubB); entorhinal cortex (EC); fusiform gyrus (FG).

### Higher p-tau burden in parahippocampal regions in Aβ-high versus Aβ-low centenarians

These observations prompted us to further investigate subregional p-tau burden in the context of low versus high Aβ. To capture the stage at which Aβ and p-tau interactions are most dynamic, and to account for higher Braak stages in Aβ-high centenarians (**Figure S1**), which are associated with higher p-tau burdens (**Figure 3A**), we restricted the analysis to 82 centenarians with Braak stages III–IV. Compared to 38 Aβ-low centenarians, the 44 Aβ-high centenarians had higher p-tau burdens in the parahippocampal subregions presubiculum-A and -B, entorhinal cortex (EC), and fusiform gyrus cortex (FG), while p-tau burden in hippocampal subregions did not differ (**Figure 4**). A similar trend was observed between 11 Aβ-high and 11 Aβ-low centenarians with Braak stages I-II (**Figure S4**), suggesting an early role for Aβ in influencing parahippocampal p-tau burden. This was also reflected in strong and consistent correlations between Aβ (burden and Thal phase) and parahippocampal p-tau burden, while correlations between Aβ and hippocampal p-tau burden were limited (**Figure 3A**).

**Figure 4.**
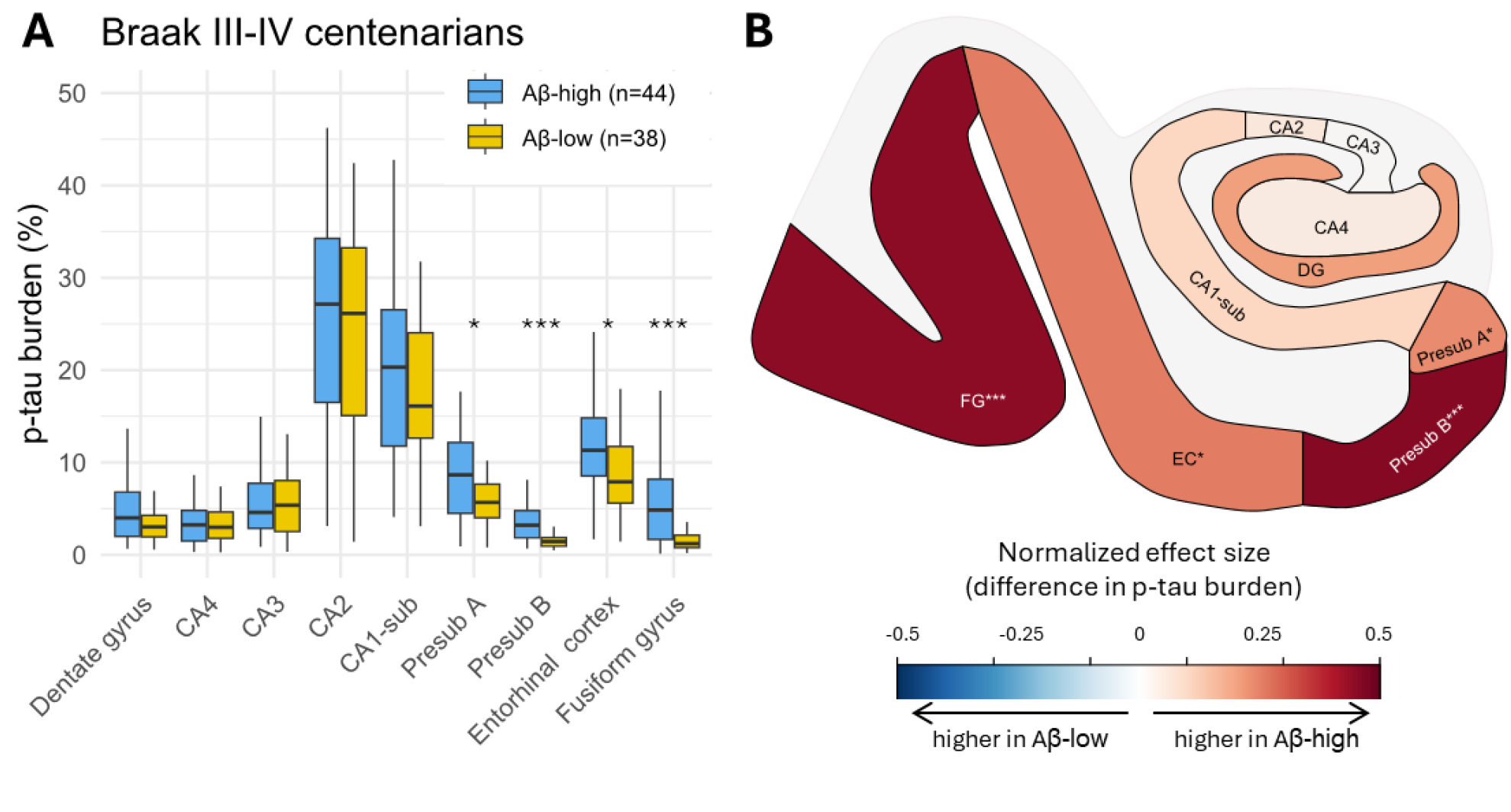
Higher parahippocampal p-tau burden in Aβ-high versus Aβ-low centenarians with Braak stages III-IV. **(A)** Comparison of p-tau burden (%) across nine subregions in the medial temporal lobe in 44 Aβ-high centenarians with intermediate Alzheimer’s disease neuropathologic change (ADNC) and 38 Aβ-low centenarians with definite or possible primary age-related tauopathy (PART), all with Braak stage III-IV (see Figure 1). Subregional p-tau burdens in ADNC and PART groups were compared with Wilcoxon rank-sum tests (p* ≤0.05 and ** ≤0.01, ***≤≤***≤0.001).**(B)** The magnitude and direction of the difference in subregional p-tau burden between Aβ-high and Aβ-low centenarians was calculated as a normalized effect size, which ranged from -1 (p-tau burden higher in Aβ^-^) to 1 (higher in Aβ-high).

### Half of Aβ-high centenarians is resistant to high FG p-tau burden typical for AD

Notably, higher p-tau burden in the parahippocampal subregions also associated with a lower CA2/CA1 p-tau burden ratio (Figure 3A), which suggests two distinct patterns of tau distribution in which centenarians have either a high ‘PART-like’ CA2/CA1 p-tau burden ratio *or* a high ‘AD-like’ parahippocampal p-tau burden. This was reflected in the independent reference-cohorts, where thresholds for CA2/CA1 p-tau burden ratio (>1.6) and FG p-tau burden (<4%) entirely separated the reference Aβ-low PART-cases from the Aβ-high AD-patients (**Figure 5A**), suggesting mutual exclusivity. Applying these thresholds to 107 centenarians (FG p-tau burden was unavailable for 4 Aβ-low centenarians and 1 Aβ-high centenarian) indicated that 39% of centenarians were resistant to p-tau accumulation typical for AD or PART (CA2/CA1 ratio <1.6 and FG <4%; 42/107), suggesting that these centenarians may have an age-related related p-tau distribution separate from typical PART and AD. Furthermore, 29% had a ‘PART-like’ distribution (CA2/CA1 ratio >1.6 and FG <4%; 31/107), 23% had an ‘AD-like’ p-tau distribution (CA2/CA1 ratio <1.6 and FG >4%; 25/107), and 8% had a ‘mixed’ p-tau hippocampal distribution (CA2/CA1 ratio >1.6 and FG >4%; 9/107) (**Figure 5B**). Representative images for each of these four p-tau distribution patterns are shown in **Figure 5C**. ‘AD-like’ and ‘mixed’ p-tau distributions were (almost) exclusively observed in Aβ-high centenarians. However, only half of Aβ-high centenarians had an ‘AD-like’ p-tau distribution (31/62), while the other half exhibited resistance to high FG p-tau burden and displayed either a ‘PART-like’ distribution (n=19) or p-tau resistance (n=12; **Figure 5D**). Notably, Aβ burden was slightly higher in many subregions in those Aβ-high centenarians with ‘AD-like’ or ‘mixed’ p-tau distribution, suggesting that not merely presence of Aβ, but also the level of Aβ burden has an influence on p-tau distribution (**Figure S5**).

**Figure 5.**
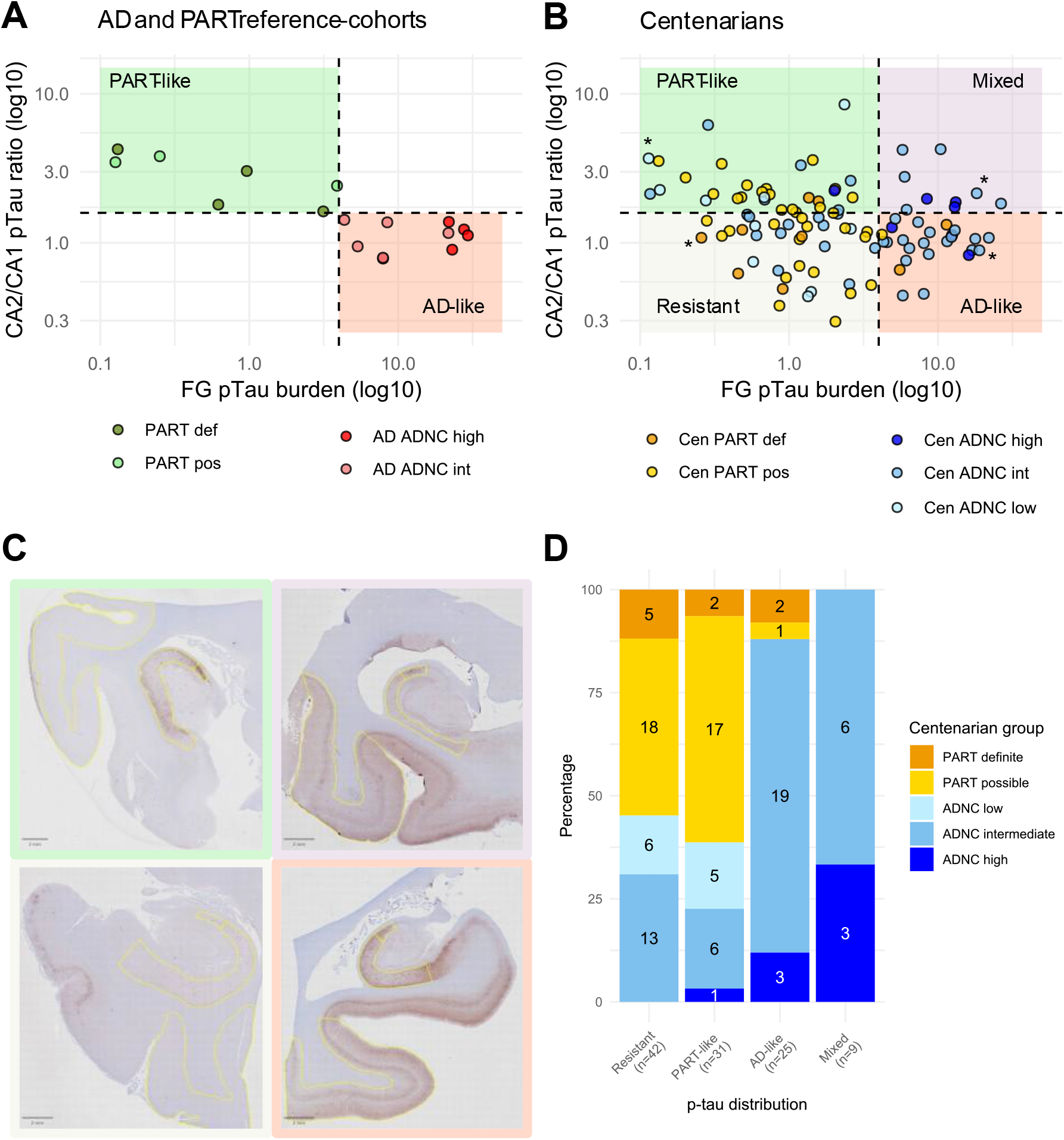
Four different p-tau distributions in the medial temporal lobe. **(A)** PART and AD cases from the reference cohorts could be separated based on thresholds for the CA2/CA1 p-tau burden ratio (1.6) and p-tau burden in the fusiform gyrus (FG; 4%). **(B)** This generated a quadrant with ‘PART-like’, ‘AD-like’, ‘Resistant’, and ‘Mixed’ p-tau distributions in which the centenarians could be categorized (n=107;, because FG p-tau burden was missing for 4 Aβ-low and 1 Aβ-high centenarians). Axis on a log 10 scale to improve visibility of cases with a low FG burden. **(C)** Examples of p-tau distribution of the most characteristic centenarian in every quadrant, as determined by the Euclidean distance, and marked with an asterisk in (B). Subregions CA2, CA1, and FG are outlined in yellow. **(D)** Aβ-high and Aβ-low centenarians (i.e., meeting the criteria for ADNC or PART, see Figure 1) were differentially distributed across the four p-tau distributions (X2=49.185, df=12, p<0.001).

### Centenarians who are resistant to accumulating Aβ-related p-tau burden maintained cognition

Cognitive performance did not differ significantly between Aβ-low and Aβ-high centenarians (median *z-*scored global cognition 0.06 versus -0.11; p= 0.335; **Figure 6A**). However, performance on memory, executive functioning, and global cognition was highest in centenarians with ‘PART-like’ and ‘resistant’ p-tau distributions (median *z-*scored global cognition 0.60 and 0.04, respectively), while performance was lower in centenarians with Aβ-related ‘AD-like’ or ‘mixed’ p-tau distributions (−0.48 and -0.59, respectively; p=0.004; **Figure 6B**). Subsequent explorative analysis showed that Aβ-high centenarians with a low FG p-tau burden (i.e., ‘PART-like’ or ‘resistant’ p-tau distributions, n=31) performed better than Aβ-high centenarians with a high FG p-tau burden (i.e., ‘AD-like’ or ‘mixed’ p-tau distributions, n=31) on memory (median z-scored performance 0.32 versus -0.60, p=0.002), fluency (0.20 versus -0.32, p=0.013), executive functioning (0.32 versus -0.43, p=0.003), and global cognitive functioning (0.37 versus -0.51, p=0.002; **Figure 6C**). This implies that the presence of Aβ is required for higher p-tau burden in the parahippocampal subregions, and that it is the p-tau burden and not the presence of Aβ itself that leads to associations with lower cognitive performance.

**Figure 6.**
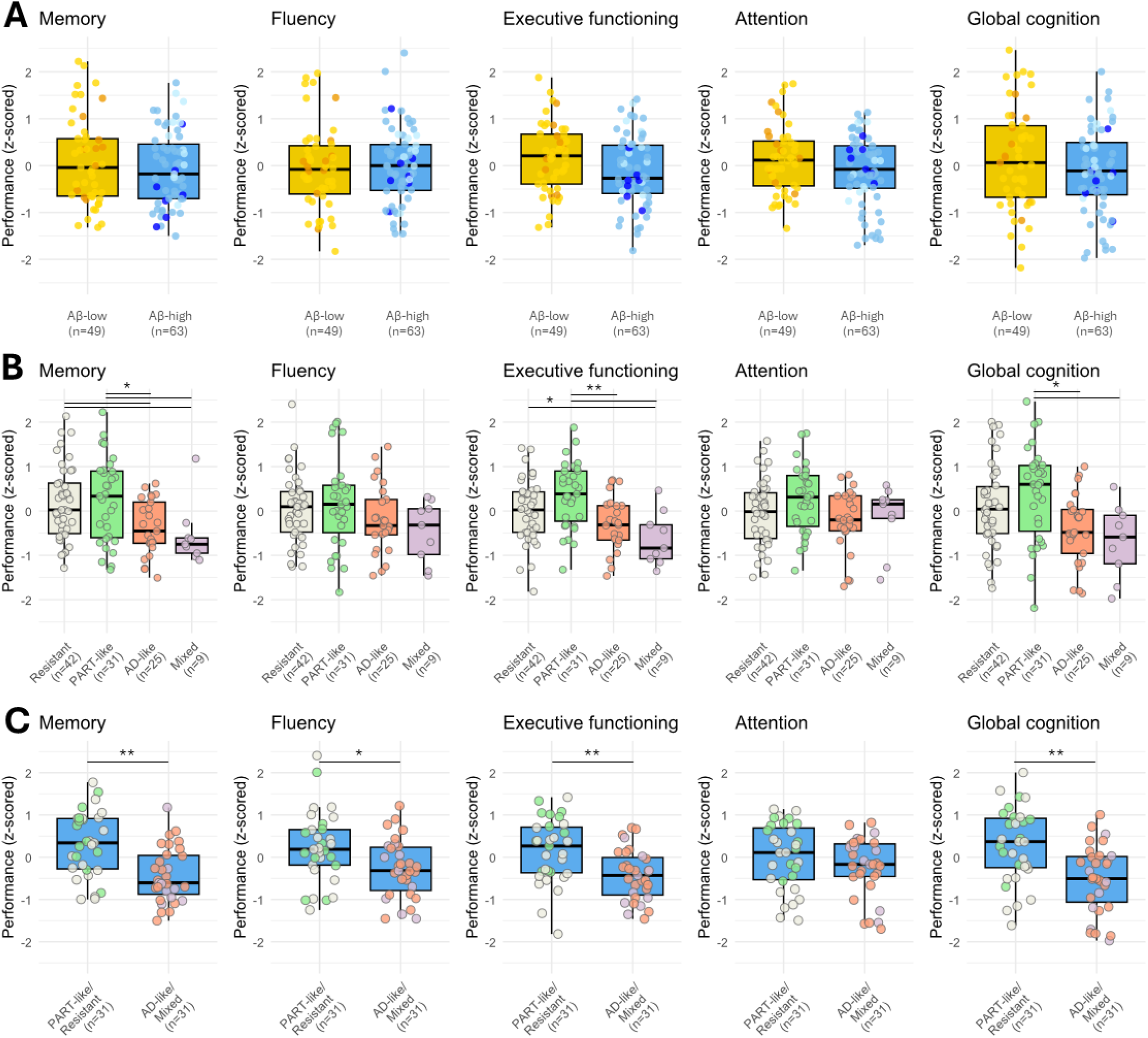
Lower cognitive performance in centenarians with AD-like and mixed p-tau distribution. **(A)** Comparison of cognitive performance on different domains and global cognition between centenarians grouped as Aβ^-^-low (i.e., Thal phase ≤2/primary age-related tauopathy (PART)) or Aβ-high (i.e., Thal phase ≥3/Alzheimer’s disease neuropathologic change (ADNC; low, intermediate or high)). Individual points indicate definite PART (orange, n=10), possible PART (yellow, n=39), low ADNC (light blue, n=11), intermediate ADNC (blue, n=45) high ADNC (dark blue, n=7). **(B)** Comparison of cognitive performance between centenarians with four different p-tau distributions based on the CA2/CA1 p-tau burden ratio and FG p-tau burden: Resistant; PART-like; AD-like; mixed (Figure 5). **(C)** Comparison of cognitive performance in 61 Aβ-high centenarians with an PART-like/Resistant p-tau distribution (i.e., FG p-tau burden < 4%, n=31) or an AD-like/Mixed p-tau distribution (i.e., FG p-tau burden >4% n=31). Group differences were assessed using Wilcoxon rank-sum tests (A and C) or Kruskal-Wallis rank sum test (B), followed by Dunn’s post-hoc test corrected for FDR. p (* ≤0.05 and ** ≤0.01).

To further evaluate the independent associations of subregional Aβ and p-tau burdens with cognitive performance, we assessed associations between subregional Aβ burden and cognition while adjusting for p-tau burden in the same subregions, and vice versa. Interestingly, subregional Aβ burden (corrected for subregional p-tau burden) did not associate with cognitive performance, except for some weak associations between higher Aβ burden in the CA2 and lower memory, executive functioning, and global cognition (**Figure 7A**, **Table S2**). In contrast, increased p-tau burden in the parahippocampal regions presubiculum-A and -B, EC, and FG (the latter used to define ‘AD-like’ and ‘mixed’ p-tau distributions) independently associated with lower performance on memory, executive functioning, and global cognitive functioning (**Figure 7B**, **Table S2**). Moreover, cognitive performance was not associated with p-tau burden in the hippocampal subregions CA2, CA3, CA4, or DG, nor with the PART-associated CA2/CA1 p-tau burden.

**Figure 7.**
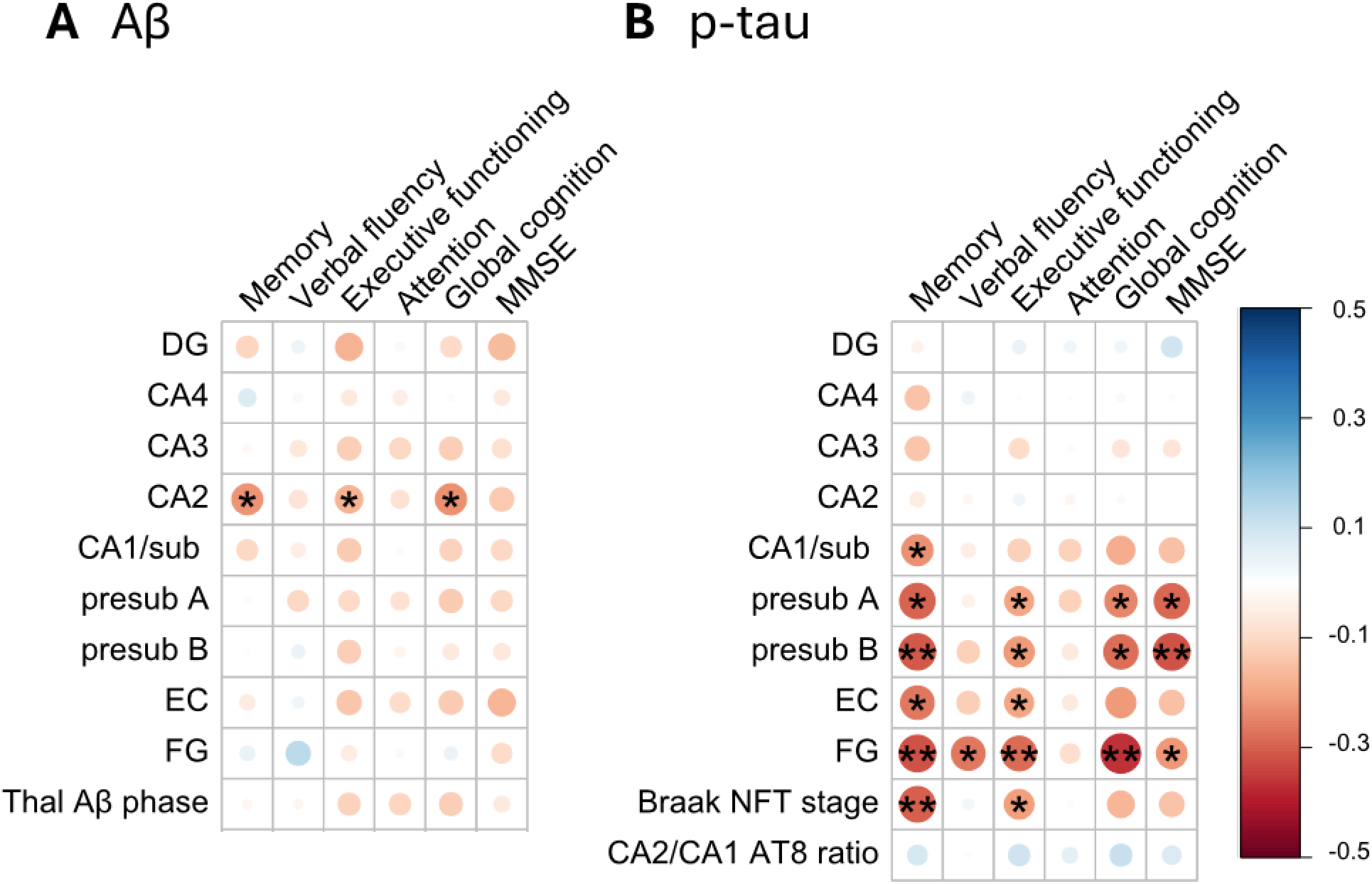
Higher p-tau, and not Aβ, in the parahippocampal subregions associates with lower cognitive performance. Robust linear regression analysis between cognitive performance in 112 centenarians and (A) Aβ and (B) p-tau burden in 9 subregions of the medial temporal lobe (MTL): dentate gyrus (DG); cornu ammonis (CA)4-2; CA1-subiculum (CA1/sub); presubiculum A and B (presub A; presub B); entorhinal cortex (EC); fusiform gyrus (FG). Color of the circles indicate the direction and strength of the regression coefficient, exact regression coefficients are given in Table S2. Models were corrected for covariates of age at death, sex, years of education, TDP-43 stage (0-3) and hippocampal sclerosis. Moreover, Aβ models were corrected for the corresponding subregional p-tau burden, and p-tau models were adjusted for the corresponding subregional Aβ burden. Thal Aβ phase was corrected for Braak NFT stage, and vice versa. The CA2/CA1 p-tau burden ratio was not corrected for Aβ. All variables were standardized (z-scored) to ensure comparability between the regression coefficients. P-values were FDR corrected with * p≤0.05, ** p≤0.01, and *** p≤0.001), and can be found in Table S2.

### Decoupling of Aβ spread and burden reveals resilience mechanisms in centenarians

As both the ADNC and PART criteria include Thal phase to distinguish low from high Aβ pathology^13,16^, we classified centenarians as Aβ-low or Aβ-high accordingly (**Figure 1A**). However, we previously reported that a subset of centenarians with Thal phase ≥3 exhibited discordantly low neocortical Aβ burden while maintaining cognitive performance^7^. Therefore, we explored the differential relation between Thal phase (i.e., Aβ spread) and Aβ burden on p-tau burden in the FG and cognition. For this, we applied a threshold representing the minimal Aβ and p-tau burdens observed in the FG in the independent AD cohort (1.2% and 4%, respectively), which segregated the independent AD and PART cohorts completely (**Figure 8A**).

**Figure 8.**
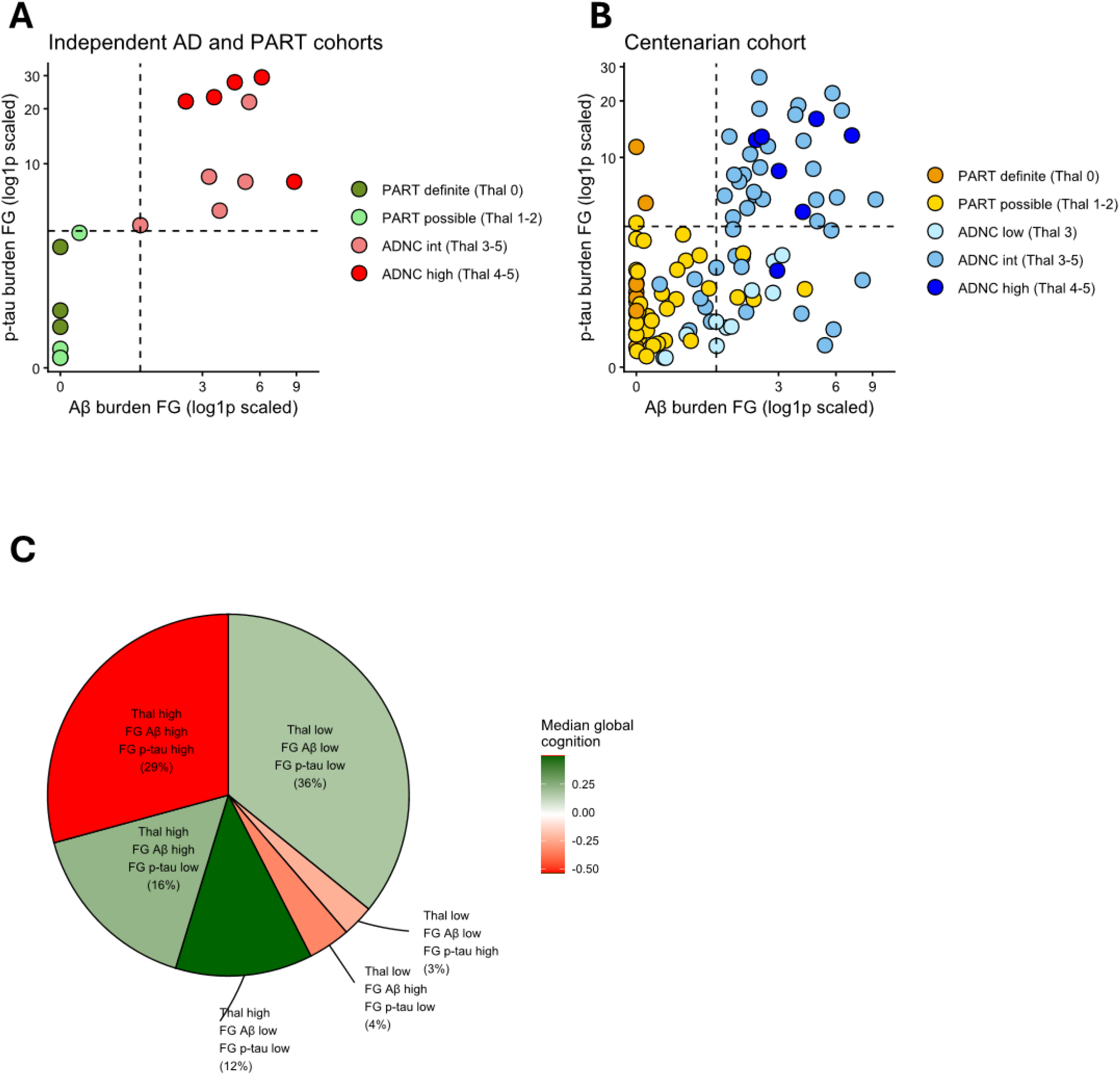
Heterogeneous relationships between Aβ spread, Aβ burden, p-tau, and cognition in centenarians. **(A)** Independent AD and PART reference cohorts are fully separated using thresholds for fusiform gyrus (FG) Aβ burden (1.2%) and FG p-tau burden (4%), indicated by the dashed lines. Colors indicate diagnostic groups based on Alzheimer’s Disease Neuropathologic Change (ADNC; low, intermediate (int) and high) and primary age-related tauopathy (PART; definite or possible) criteria. The Thal phases for the spatiotemporal spread of Aβ are indicated. **(B)** Application of these thresholds to the centenarian cohort (n=106 for which all three measurements were available) shows a heterogeneous distribution across Aβ burden and p-tau burden, despite similar classification by ADNC and PART criteria. **(C)** Pie chart summarizing the proportion of centenarians within each quadrant defined by low or high Thal phase (≤2 or ≥3, respectively), FG Aβ burden (low/high), and FG p-tau burden (low/high). Color intensity reflects median global cognitive performance per group, with positive values in green representing better cognition.

Centenarians showed a more heterogenous relationship between Aβ spread, Aβ burden, and p-tau burden compared to reference AD and PART cases. The largest group of centenarians (36%) had low Aβ-spread (i.e., Thal phase ≤2), low FG Aβ burden, and low FG p-tau burden, resembling the PART cases from the independent reference cohort (**Figure 8B**). The median z-normalized global cognitive performance in this group was high (0.17; **Figure 8C**). On the other end of the spectrum, the second largest group of centenarians (29%) had high Aβ-spread, high FG-Aβ burden, and high FG p-tau burden, resembling the AD cases from the independent reference cohort. The median global cognitive performance in this group was low (−0.54). Interestingly, 12% of centenarians had high Aβ-spread, classifying them as ADNC, but maintained low FG Aβ and p-tau burden (the blue dots in the lower left quadrant in **Figure 8B**). The median cognition of this group was high (0.50). Another group of centenarians (16%) had high Aβ-spread and FG Aβ burden, but low p-tau burden (the blue dots in the lower right quadrant), and their global cognitive performance also remained high (0.22). Altogether, these findings suggest that centenarians may maintain cognitive performance through three successive stages of resilience along the amyloid-cascade: (1) by resisting advanced Aβ spread beyond Thal phase 2 and consequently remaining within the PART spectrum, with limited p-tau accumulation (36%); (2) among those with advanced Aβ spread, by being resilient to the subsequent accumulation of high Aβ burden (12%); and (3) among those with both advanced Aβ spread and high Aβ burden, by being resilient to the downstream accumulation of a high p-tau burden (16%). At the end of this cascade, transitioning from resilience into affected, is a group of centenarians who exhibit high Aβ spread, high Aβ burden and high p-tau accumulation and show lower global cognitive performance.

## Discussion

We investigated whether centenarians have an altered relationship between Aβ pathology and p-tau distribution in the MTL, and whether this relates to maintained cognitive performance, as this could point to naturally occurring protective mechanisms that preserve a healthy functioning brain until extreme old age. We observed that Aβ pathology in the MTL was required for higher parahippocampal p-tau burdens to accumulate, and that p-tau burden, more than Aβ burden itself, was associated with cognitive decline. Strikingly, approximately half of the centenarians with Aβ pathology appeared resilient to the Aβ-driven progression of p-tau into the parahippocampus and maintained high levels of cognitive performance. Together, these findings suggest that centenarians who preserved cognition until extreme age did so either by resisting Aβ pathology altogether, or by being resilient to Aβ-driven p-tau progression.

Emerging evidence suggests that this resistance to Aβ pathology or it’s decoupling from tau propagation may be partly genetically mediated. First of all, we reported that the *APOE* ε2 allele is enriched in centenarians and strongly associates with resistance to Aβ pathology^7,17^. Moreover, we recently characterized a *TMEM106B* haplotype that is enriched in centenarians and specifically protects against the progressive accumulation of p-tau (Salazar *et al.,* 2026a and 2026b, submitted to Nature Medicine). Furthermore, mechanisms underlying resilience in centenarians may overlap with those observed in *PSEN1* mutation carriers who carry the protective *APOE3* Christchurch or *RELN-COLBOS* mutations and developed extensive Aβ pathology but atypical low levels of p-tau pathology^18,19^. Together, these independent observations, spanning both extreme aging and autosomal dominant AD in much younger individuals further strengthen support for the broader existence of biological resilience mechanisms that uncouple Aβ pathology from downstream tau propagation.

This interpretation is consistent with proteomic analyses of the medial temporal gyrus in a subset of this cohort, which revealed that all centenarians maintained low local MTBR-tau peptide abundance despite in some cases substantial Aβ abundance (Hulsman et al. 2026). These centenarians exhibited preserved proteostasis, metabolic adaptation, lower ApoE abundance, and maintained lipid β-oxidation (Hulsman *et al*., 2026, submitted to Nature Medicine). Another recent work including centenarians from this cohort pointed to transitions in microglial state at the Aβ-tau interface as mechanisms underlying resilience^20^, while others have suggested the maintenance of the excitatory-inhibitory balance as a potential characteristic of natural neuronal resilience to AD neuropathology^21^. All these possible mechanisms may contribute to resistance and resilience to Aβ pathology and warrant further investigation.

We previously reported that higher neocortical Aβ burden associated with lower cognition in this cohort of centenarians^7^. Here we show that the association between Aβ burden in the MTL and cognition weakened substantially after additional correction for p-tau burden. This aligns with the concept that neocortical Aβ mediates propagation of p-tau from the MTL into the neocortical areas via the FG, where p-tau burden rather than Aβ itself drives cognitive decline^22,23^. Whether neocortical Aβ burden independently affects cognitive performance in this group of centenarians, or whether this association is partly mediated by neocortical p-tau burden, remains unclear. However, given the low number of centenarians with substantial neocortical p-tau pathology (only 8/112 had Braak stage V), it is unlikely that the remaining association after correction for Braak stage is completely mediated by neocortical p-tau burden7. Moreover, further research is needed to investigate the contribution of co-pathologies such as TDP-43 and Lewy Bodies on the relationship between Aβ, p-tau propagation and cognition. Notably, 12% of centenarians had low Aβ burden while having advanced Aβ spreading (Thal≥3), which supports a model in which Aβ spreading precedes increases in local Aβ abundance. We cannot exclude that, had these individuals lived longer, Aβ levels might eventually have exceeded a threshold level that facilitates tau propagation and affects cognition.

All 112 centenarians investigated in this study had some degree of p-tau pathology, suggesting that p-tau accumulation is an inevitable consequence of reaching extreme old age. However, this does not imply inevitable cognitive decline: the largest group of centenarians (39%) exhibited a distribution of p-tau pathology that was distinct from elderly affected with PART- or AD-related dementia (i.e., low CA2/CA1 p-tau burden ratio and low FG p-tau burden). Interestingly, a similar non-specific p-tau distribution was observed in a 115-year-old Dutch woman who maintained exceptional cognition until death despite neuropathological features consistent with PART (i.e., Thal phase 0 and Braak stage II)^24^. Together, these observations raise the possibility that some patterns of p-tau accumulation at extreme old age are not invariably a marker of neurodegenerative disease, but instead may reflects a potentially benign age-related phenomenon distinct from the pathogenic mechanisms underlying PART and AD.

The second largest group of centenarians (29%) had a PART-related p-tau distribution (i.e., a high CA2/CA1 p-tau burden ratio and low FG p-tau), which also did not associate with lower cognitive performance. While the CA2/CA1 p-tau burden ratio was previously described to differentiate between AD and PART^9,10^ and showed the same pattern in our reference cohorts, it did not distinguish between Aβ-high and Aβ-low centenarians. Notably, one-third of Aβ-high centenarians demonstrated a high CA2/CA1 p-tau burden ratio, suggesting that PART-like p-tau distributions may coexist with AD-related pathology. While this aligns with the established concept that the aged brain often presents with mixed rather than uniform pathologies^2,3,6^ it challenges current diagnostic frameworks that treat PART and ADNC as mutually exclusive entities.^13,25^ Our findings therefore support recently proposed revisions of the criteria for PART, in which low levels of ADNC and (possible) PART may occur simultaneously in the same brain.^14^ Importantly, PART-like p-tau distributions were not associated with cognitive performance, regardless of Aβ status, highlighting the clinical value of classifying p-tau distribution independently of Aβ pathology.

While our AD and PART cohorts provided valuable reference points, larger sample sizes, such as those investigated in the PART Working Group^9^ are needed to better capture the heterogeneity of p-tau spatiotemporal patterns in AD and PART cases and relationships with age. Such studies may help systematically identify key determinants of PART-like, AD-related, and age-related distributions of p-tau. Finally, we quantified total Aβ and p-tau burden using widely applied antibodies (6F/3D and AT8)^26,27^. Future studies examining specific isoforms (e.g., Aβ_40_, Aβ_42_, Aβ_N3pE_, Aβ_pSer8_, 3R/4R tau),^13,28,29^ distinct aggregation states, and stages of neurofibrillary tangle maturation may further refine our understanding of mechanisms that protect against cognitive decline into extreme old age.^29,30^

Our findings suggest that the amyloid cascade hypothesis may require refinement, as Aβ accumulation alone is not sufficient to drive tau pathology but depends on individual susceptibility or resilience to Aβ-associated p-tau propagation. The identification of resistance and resilience to Aβ as distinct mechanisms of protection provides an opportunity to explore endogenous pathways that preserve cognitive health into extreme age. While the first possible genetic and neurobiological drivers are being uncovered, both warrant further exploration, as they may offer complementary therapeutic entry-points: preventing the accumulation of Aβ pathology altogether, or limiting downstream p-tau propagation once Aβ pathology has already emerged.

## Methods

### The 100-plus Study

The 100-plus Study follows centenarians who self-report to be cognitively healthy at study-inclusion.^5^ At baseline and annual follow-up house-visits we collect lifetime history including educational history, assess cognitive performance, and discuss optional post-mortem brain donation. The study was initiated in 2013, approved by the VUmc medical ethics committee (2016.440) and all participants provided written informed consent.

### Cognitive assessment of centenarians

Cognitive performance was annually assessed with a test battery composed of 18 (sub)tests covering memory, fluency, executive functioning, and attention (see supplement; Methods).^5^ To minimize the interval between cognitive assessment and neuropathology, we analyzed performance at the last study visit, which occurred a median of 9 months prior to death (inter quartile range (IQR): 4–14). For some centenarians, a subset of cognitive test scores were missing due to fatigue, sensory or motor difficulties, or revision of the cognitive test battery over time. To address this issue, missing test scores were imputed using multiple imputation by chained equations, as described in the supplement and in line with previous publications.^6,7,31,32^ We only included tests for which >50% of the test scores was available for the entire cohort, resulting in the inclusion of 13 tests (see supplement; Table S1). Individual test scores were z-normalized across the 112 centenarian brain donors, and domain-specific performance scores for memory, fluency, executive functioning, and attention were calculated by averaging the normalized scores (see supplement; and Table S1). Global cognition was measured with the Mini-Mental State Examination (MMSE) and by calculating a composite global cognition score by averaging the four-domain specific z-scores.

### Brain donation and tissue processing

Brain donations from centenarians, AD-patients and PART-cases (detailed two sections below) occurred between 2001 and 2024, were performed by the Netherlands Brain Bank (NBB; Amsterdam, The Netherlands), and were approved by the VUmc medical ethics committee (2009.148). Donors provided written informed consent for autopsy, tissue storage, and use of anonymized clinical and neuropathological data. Brain donation occurred within 12 hours post-mortem (median 6 hours, IQR: 5-7 hours), followed by fixation of the right hemisphere in 10% formalin for ∼4 weeks.

### Neuropathological evaluation and diagnosis of ADNC and PART in centenarians

Brains from 112 centenarian were assessed for: (A) Thal Aβ phase (0-5);^33^ (B) Braak NFT stage (0-VI);^34^ and (C) CERAD neuritic plaque (NP) score (“absent” [0], “sparse” [1], “moderate” [2], or “frequent” [3]);^35^ which were combined into an ABC score to determine the level of AD neuropathologic change (ADNC; not, low, intermediate, high)^25^ or the presence of PART (Figure 1A).^13^ Centenarians with Thal phase ≥ 3, were classified as Aβ-high (which included low, intermediate, or high ADNC), centenarians with Thal phase ≤ 2, were classified as Aβ-low (which included definite and possible PART).

### AD and PART reference cohorts

Hippocampal pathology in centenarians was compared with that observed in 11 individuals with ADNC patients and 7 individuals with PART cases with dementia from the NBB (**Table 1**). Cases were selected based on: 1) a clinical dementia diagnosis; 2) availability of formalin-fixed paraffin-embedded (FFPE) hippocampal tissue; 3) neuropathological diagnosis of intermediate/high ADNC^25^ for AD, or definite/possible PART.^13^ PART cases meeting these criteria were scarce in the NBB; therefore, cases with Lewy Body pathology (LB) and transactive response DNA binding protein 43 (TDP-43) co-pathologies were included. However, cases with severe hippocampal sclerosis (HS) were excluded, as extensive neuronal cell loss may bias p-tau measurements in the CA1 subfield. The primary clinical diagnosis of the reference PART cohort was either ‘tangle-only dementia’ (n=2) or Parkinson’s disease dementia (n=5).

### Subregional Aβ and p-tau burden

FFPE sections of the right MTL including the hippocampus at the level of the lateral geniculate nucleus were cut at 6µm. Immunohistochemistry was applied to visualize Aβ (clone 6F/3D, DAKO, M0872) and p-tau (AT8, Thermo Scientific, MN1020; detailed protocol in supplementary material). Stained sections were scanned at 20X magnification with an Olympus VS200 slide scanner and software (v3.3), and analyzed in QuPath (v0.4.2).^36^ Consistent with previous studies,^7,10,37–39^ the MTL was segmented in five hippocampal and four parahippocampal subregions: dentate gyrus (DG), Cornu Ammonis (CA)4, CA3, CA2, CA1/subiculum, presubiculum part A and B (presub A/B), entorhinal cortex (EC), and fusiform gyrus cortex (FG) (detailed segmentation protocol in supplementary material). A pixel classifier was trained to detect Aβ and p-tau pathology and calculate the pathology burden as the percentage of a subregion positive for Aβ or p-tau.^7^ The CA2/CA1 p-tau burden ratio was calculated by dividing p-tau burden in the CA2 subregion by the burden in CA1/subiculum, as previously described.^10^ To ensure that the calculated CA2/CA1 p-tau burden ratio s reflected actual p-tau distributions and were not affected by near-zero noise, all cases with p-tau burden <1% in CA1/subiculum or CA2 (n=2) were visually inspected and confirmed to have sufficient staining for reliable ratio calculation.

### Statistical analysis

Analyses were performed in RStudio (v4.3.2). Comparisons between two-groups were made with Wilcoxon rank-sum tests, and multiple-group comparisons were conducted with Kruskal–Wallis tests followed by Dunn’s post hoc tests (dunn.test, v1.3.6). P-values were corrected for multiple testing using the Benjamini–Hochberg false discovery rate (FDR) method. Subregional p-tau burden was compared between Aβ-high and Aβ-low centenarians using Wilcoxon rank-sum tests, with normalized effect sizes calculated via wilcox_effsize from the rstatix package (v0.7.2). Effect sizes ranged from -1 (higher in Aβ-low centenarians) to 1 (higher in Aβ-high centenarians) and were visualized using a custom-made R-based hippocampal illustration. Spearman correlations were used to assess associations between Aβ and p-tau burdens.

Associations between subregional Aβ and p-tau burden and cognitive scores were examined using robust linear regression models (MASS v7.3-60) to reduce outlier impact and address non-normal residuals, with estimated p-values extracted from t-values (sfsmisc v1.1.19). Models were adjusted for the co-variates sex, age at death and education (years), and of TDP-43 stage (0-3)^40^, and hippocampal sclerosis (absent or present), previously shown to associate with cognitive performance in centenarians.^31,41^ To investigate the independent effects of Aβ and p-tau burden on cognitive scores, Aβ models were adjusted for the corresponding subregional p-tau burden, and p-tau models for the corresponding subregional Aβ burden.

All variables were normalized to *z*-scores to ensure comparability of regression coefficients across different associations. Because performance on the different cognitive domains was highly intercorrelated, as well as the level of different pathologies was intercorrelated, treating all tests as independent would yield overly conservative FDR correction and reduced power. We therefore estimated the effective number of independent tests (Mₑff) using the Li & Ji method (poolr v1.1-1). All p-values were subsequently pooled and adjusted for FDR and scaled by Mₑ_ff_. For all abovementioned analysis, a p-value <0.05 was considered significant.

## Data Availability

All data produced in the present work are contained in the manuscript.

## Acknowledgements

We thank and acknowledge all participating centenarians and their family members. Moreover, we would like to thank and acknowledge all other brain donors and the Netherlands Brain Bank staff for the great cooperation. Furthermore, we would like to acknowledge all current and former practical team members of the 100-plus Study that contributed to data collection, and Andrea B. Ganz, PhD for previous contributions to neuropathological evaluations. Moreover, we thank Linda M. C. Lorenz for her contributions to interpreting of neuropsychological data. We would also like to thank Sanne M.M. Vermorgen for her assistance with selecting PART cases from the Netherlands Brain Bank.

## Funding

### Author contributions

The 100-plus Study was initiated and designed by H.H. Neuropathological evaluation was performed by A.J.M.R. and the NBB, supported by S.K.R. and M.C.L. Immunohistochemistry was performed by S.K.R. and M.C.L. Quantitative pathology analyses were performed by S.K.R., under supervision of T.E.R. and J.M.W. Design and interpretation of the neurocognitive testing procedures of the 100-plus Study was supported by S.A.M.S. Imputation of missing test scores was performed by M.H., and verified by S.K.R. and S.A.M.S. Data analyses were performed by S.K.R., supported by M.H. A Custom-made digital hippocampus illustrations were designed by S.J.vd.L. The research was supervised by T.E.R., J.J.M.H., J.M.W., and H.H. Design of the research involved S.K.R., T.E.R., J.J.M.H., J.M.W., and H.H. All authors commented on previous versions of the manuscript and approved the final manuscript.

## Notes

### Competing Interest Statement

The authors have declared no competing interest.

### Author Declarations

The VUmc Medical Ethics committe gave ethical approval for this work (2009.148 and 2016.44).

### Funding

- Alzheimer Nederland: WE.08-2023-03
- BrightFocus Foundation: A2021031S
- SURFsara (Netherlands): 2022.031

